# Observed strong pervasive positive selection in the N-terminal domain, receptor-binding domain and furin-cleavage sites of SARS-CoV-2 Spike protein sampled from Zimbabwean COVID-19 patients

**DOI:** 10.1101/2022.04.27.22274357

**Authors:** Milton S Kambarami, Manasa Justen, Mushiri Tawanda

## Abstract

Mutations primarily in the Spike (S) gene resulted in the emergence of many SARS-CoV-2 variants like Alpha, Beta, Delta and Omicron variants. This has also caused a number of COVID-19 pandemic waves which have impacted human lives in different ways due to restriction measures put in place to curb the spread of the virus. In this study, evolutionary patterns found in SARS-CoV-2 sequences of samples collected from Zimbabwean COVID-19 patients were investigated. High coverage SARS-CoV-2 whole genome sequences were downloaded from the GISAID database along with the GISAID S gene reference sequence. Biopython, NumPy and Pandas Data Science packages were used to load, slice and clean whole genome sequences outputting a fasta file with approximate Spike (S) gene sequences. Alignment of sliced dataset with GISAID reference sequence was done using Jalview 2.11.1.3 to find exact sequences of SARS-CoV-2 S gene. Evidence of recombination signals was investigated using RDP 4.1 and pervasive selection in the S gene was investigated using FUBAR algorithm hosted on the Datamonkey webserver. Matplotlib and Seaborn Python packages were used for Data Visualisation. A plot of Bayes factor hypothesizing non-synonymous substitution being greater than synonymous substitution (β > α) in the S protein sites showed 3 peaks with evidence of strong divergence. These 3 diverging S protein sites were found to be D142G, D614G and P681R. No evidence of recombination was detected by 9 methods of RDP which use different approaches to detect recombination signals. This study is useful in guiding drug, vaccine and diagnostic innovations toward better control of the pandemic. Additionally, this study can guide other non-biological interventions as we better understand the changes in various viral characteristics driven by the observed evolutionary patterns.

## 1.0: Introduction

A pneumonia Coronavirus disease later named Coronavirus Diseases 2019 (COVID-19) was reported in December 2019 in Wuhan China (Zhou *et al*., 2020; Chan *et al*., 2020). According to World Health Organisation (WHO) there has been 386 548 962 Global cumulative cases and 230 012 active cases in Zimbabwe as of 31 January 2022. The causative agent later named Severe Acute Respiratory Syndrome Coronavirus 2 (SARS-CoV-2) has very close genomic resemblance to a number of Coronaviruses infecting Bats and Pangolins (Chen, Liu and Guo, 2020; Leitner and Kumar, 2020; Liu *et al*., 2020). The Spike (S) protein which facilitates entry of SARS-CoV-2 into human host cells through Angiotensin-converting enzyme 2 (ACE2) receptors has been a widely researched region as a target for drugs and vaccines (Amaral *et al*., 2013; Dagotto, Yu and Barouch, 2020; Koirala *et al*., 2020).

### 1.1 Background

Viruses primarily evolve through recombination and substitution events resulting in new lineages, subtypes or strains (Arenas, 2022). A number of SARS-CoV-2 variants have emerged during the past 2 years with some resulting in elevated transmissibility and intensity of observed patient symptoms (Butowt, Bilinska and Bartheld, 2020; Eaaswarkhanth, Madhoun A A and Al-Mulla, 2020). This has led to a proposed nomenclature for naming SARS-CoV-2 lineages by Rambaut *et al*., 2020. WHO proposed a laymen nomenclature using Greek letters for variants/lineages that posed risk to global health and for easy communication between researchers and general citizens.

A study about COVID-19 and SARS-CoV-2 in Zimbabwean patients has been done by Mashe *et al*. 2021, however the study was more focussed on the epidemiology of COVID-19. In this study, the authors focussed on the Evolutionary virology of the SARS-CoV-2 S region sampled from Zimbabwean patients.

In Africa, countries like Zambia, Uganda and South Africa have studied the Phylogenetics and distribution of SARS-CoV-2 lineages in their countries (Bugembe *et al*., 2020; Simulundu *et al*., 2020; Tegally *et al*., 2020). Viral studies can be useful in predicting the origins and observed symptoms with respect to predominant SARS-CoV-2 lineages in the region such that drugs to alleviate symptoms and ways to deal with COVID-19 can be more specific to these regions. For example the Delta variant has been noted to infect the upper respiratory tract (Korber *et al*., 2020) which means drugs specific for the upper respiratory tract can be used to manage COVID-19.

For vaccine acquisition or development, health policies in Zimbabwe can have a guide on which vaccine is precise to the majority of Zimbabweans based on the evolutionary patterns observed in its SARS-CoV-2 sequences.

This study focussed on the evolutionary patterns of the Spike (S) gene of the sequences sampled from Zimbabwean COVID-19 patients over the past 2 years.

### 1.2: Problem statement

The SARS-CoV 2 spike protein gene is associated with high level of mutations which affects vaccine and diagnostics efficacy as well as transmissibility and possibly pathogenicity. Thus there is need to regularly monitor the evolutionary patterns in the spike protein to guide both biomedical and non-biomedical interventions.

### 1.3: Objectives of the research

1. To investigate phylogeny of SARS-CoV-2 S gene sampled from Zimbabwean COVID-19 patients.
2. To investigate evidence of recombination within the Zimbabwean SARS-CoV 2 S gene dataset.

4. To investigate divergence and/or purifying selection in the SARS-CoV 2 S gene sampled from Zimbabwean patients.

### 1.4: Justification of doing the research

The mutations observed in the Spike region of SARS-CoV-2 originating from other parts of the world show that SARS-CoV-2 is adapting in accordance to immunological patterns of such regions. However with the SARS-CoV-2 genomic data sampled from Zimbabwean patients, it is also necessary to study if there are specific evolutionary patterns of SARS-CoV-2 being driven by immunologically unique human ACE2 receptors of Zimbabwean COVID19 patients.

## 2:0 Literature Review

Coronaviruses are a group of positive-sense RNA viruses that have a large genome size of ∼30kb which are infectious. Coronaviruses have the largest known genome size and one of the most stable RNA viruses (Enjuanes, 2005).

Coronaviruses have caused outbreaks since 2003 under the flagship names Severe Acute Respiratory Syndrome (SARS) caused by Severe Acute Respiratory Syndrome coronavirus (SARS-CoV) which was mostly prevalent in China. Bats have been identified as the likely reservoir of SARS-CoV, whereas civet cats probably have served as intermediate host during the zoonotic transfer to humans (Lau et al 2005, Li et al. 2005).

Middle East Respiratory Syndrome (MERS) caused by Middle East Respiratory Syndrome coronavirus (MERS-CoV) originated from camels first isolated in June 2012 in a 60 year old Saudi Arabian male (Zaki *et al*. 2012). Besides the zoonotic coronavirus SARS CoV and MERS CoV, there has been established human coronaviruses (hCoV) of which 2 have been known since 1960s (van der Hoek, 2007 and McIntosh et al. 1967).

Coronavirus are associated mainly with respiratory, enteric, hepatic and central nervous system diseases. Human Coronaviruses are responsible for 10-20% of common colds and have been implicated in gastroenteritis, high and low respiratory tract infections, and rare cases of encephalitis. HCoVs have also been associated with infant necrotizing enterocolitis and are tentative candidates for multiple sclerosis (Enjuanes, 2005).

Laboratory findings have documented lymphopenia as the most frequent, followed by high C-reactive protein and high Aspartate transaminase (AST). Infection with COVID-19 is associated with significant morbidity especially in patients with chronical medical conditions. Data suggest that older age and comorbidities play a vital role in influencing severe disease and chronical outcomes. These data are useful to guide patient risk group management in the current epidemic. COVID-19 patients present predominantly with fever and cough which appear to be more frequent in adults than in children (Rodriguez – Morales et al. 2020).

SARS-CoV-2 being a RNA virus undergoes elevated mutations compared to DNA viruses due to the instability of the RNA structure. Such susceptibility to change paves way for evolution which results in microbial drug/vaccine resistance or adaptation to new host species (zoonosis). Zoonosis is how SARS-CoV-2 and HIV among other viruses crossed species barrier from animals like bats and chimpanzees respectively to humans (Kristian et al. 2020, Williams and Burdo,2009). SARS-CoV-2 has a ∼30% mutation frequency meaning there is a very likely chance of a mutation for every 3 replication cycles (Paccheti et al. 2020). Furthermore, the Coronaviruses genome has a ∼30% G-C content (Woo et al. 2010), the G-C content helps stabilise genomes because of the 3 hydrogen bonds formed when G and C hybridise so more energy is required to break 3 bonds than 2 bonds involved in A-T bonding.

## 3.0: Methodology

### 3.1: Sequence Data Manipulation

Zimbabwean SARS-CoV-2 whole genome sequences with high coverage were downloaded from the Global Initiative for Sharing Influenza Data (GISAID) database along with related Patient Data and the GISAID S gene Reference Sequence. The approximate S gene was extracted from whole genome of SARS-CoV-2 using Biopython, Panel Dataframes (Pandas) and Numerical python (Numpy) Data Science packages. Because the whole genome sequences had different lengths, a space of ∼100 nucleotides was allowed on both ends of the extracted spike gene sequences (Nucleotide number 21 500 -25 500 on whole genome).

The dataset was aligned using Clustal Omega Multiple sequence Alignment tool in Jalview 2.11.1.3 (Waterhouse *et al*. 2009). Redundant sequences and sequences with a higher number of unidentified nucleotide bases were removed from the dataset. Using the GISAID S gene reference sequence as a guide, flanking regions on both ends of the aligned S gene sequences were sliced out of the dataset. MEGA X (Kumar *et al*., 2018) was then used for codon alignment of the dataset using Clustal Omega, a pre-requisite process for investigating Pervasive Selection using FUBAR (Murell *et al*. 2013)

### 3.2: Distribution of SARS-CoV-2 lineages in Zimbabwe

The patient data csv file was loaded using Pandas and the lineage column was used to investigate the extent of SARS-CoV-2 lineages which infected the Zimbabwean population. A pie chart to show dates of sample collection according to COVID-19 pandemic waves was also plotted. Python Data Science packages used for patient data manipulation were Numpy and Pandas and for Data Visualisation were Seaborn and Matplotlib. The result chart was very scattered so the lineages were grouped according to WHO Greek letters for variants posing a risk to Global Health.

### 3.3: Recombination Analysis

Using a guide provided in the paper by Martin, Lemey and Posada, 2011, a number of recombination detection methods found in RDP 4.100 which use different approaches to identify recombination were selected to investigate evidence and extent of recombination in the dataset. The methods selected were RDP, GENECONV, Chimaera, MaxChi and BootScan (Padidam, Sawyer and Fauquet, 1999; Martin et al. 2005; Posada and Crandall, 2001; Smith, 1992) using default settings. A Monte Carlo-based method, SISCAN (Gibbs, Armstrong and Gibbs, 2000) was further used for thorough investigation of recombination signals in the dataset.

### 3.4: Substitution Analysis

The codon-aligned S gene dataset was uploaded onto the DataMonkey webserver for pervasive selection analysis using FUBAR. A table of values showing relationship between synonymous (α) and non – synonymous (β) rates was exported alongside the fitted Phylogenetic tree from the FUBAR Analysis. The resultant fitted Phylogenetic Tree was exported in Newick format and uploaded on iTOL webserver for plotting the unrooted tree for validation of FUBAR results. The table of values exported in csv format was used to investigate positive and negative selection in the dataset. A bar graph was plotted to investigate sites that showed strong positive selection using the Bayes Factor. The hypothesis used for Bayes factor was in assertion of evidence for pervasive positive selection (β > α). To pinpoint which sites showed strong positive selection, a threshold posterior probability of 0.9 for β > α was used to select outstanding sites. A threshold posterior probability of 0.9 for α < β was also used to investigate sites that showed strong negative selection.

## 4.0: Results

### 4.1: Distribution of SARS-CoV-2 lineages in Zimbabwe

**Figure 1:**
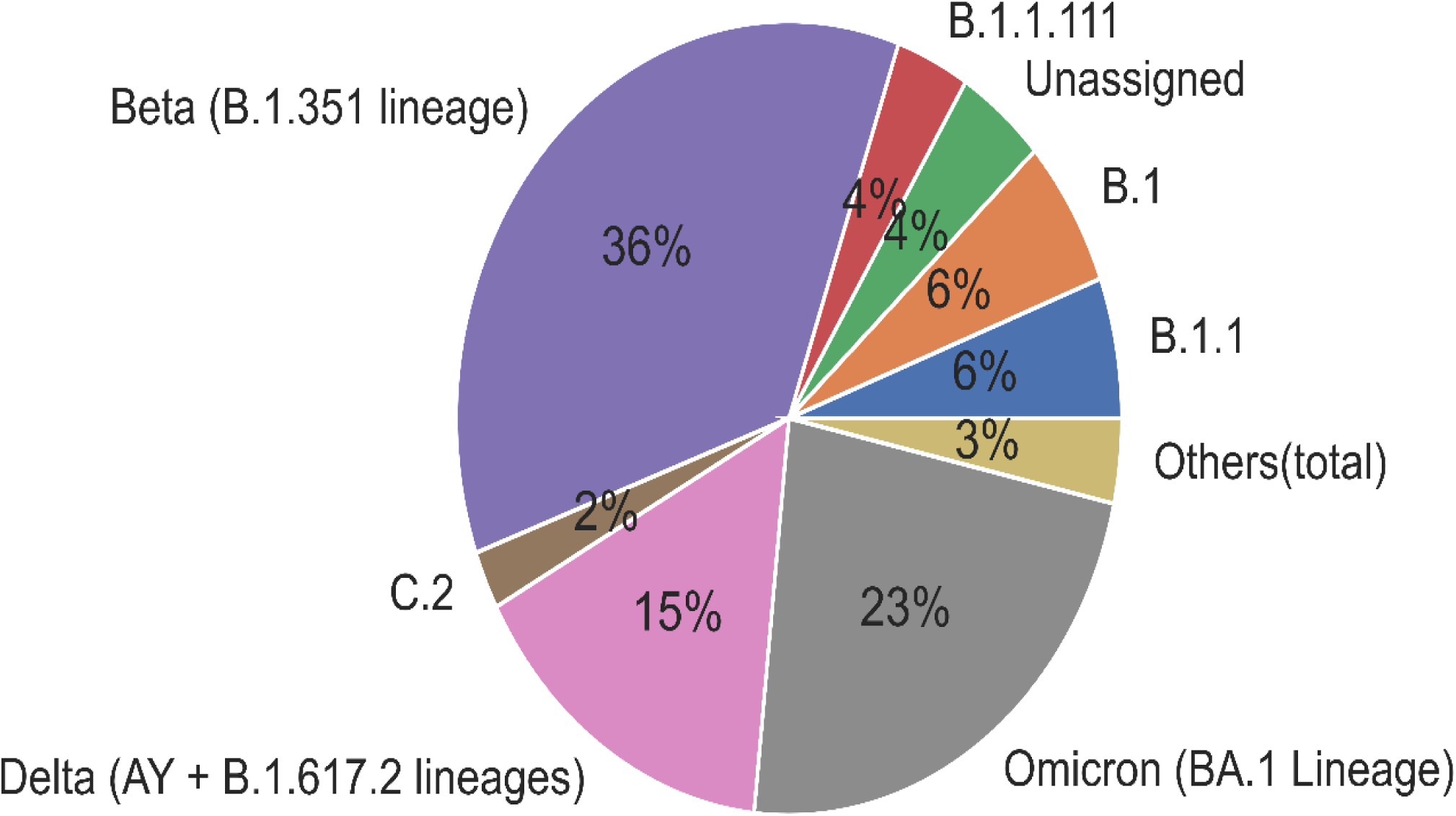
Percentages of SARS-CoV-2 lineages that were predominant over the course of the COVID-19 pandemic in Zimbabwe (up to December 2021). The Beta lineage constitutes most of the COVID-19 infection for collected samples.

**Figure 2:**
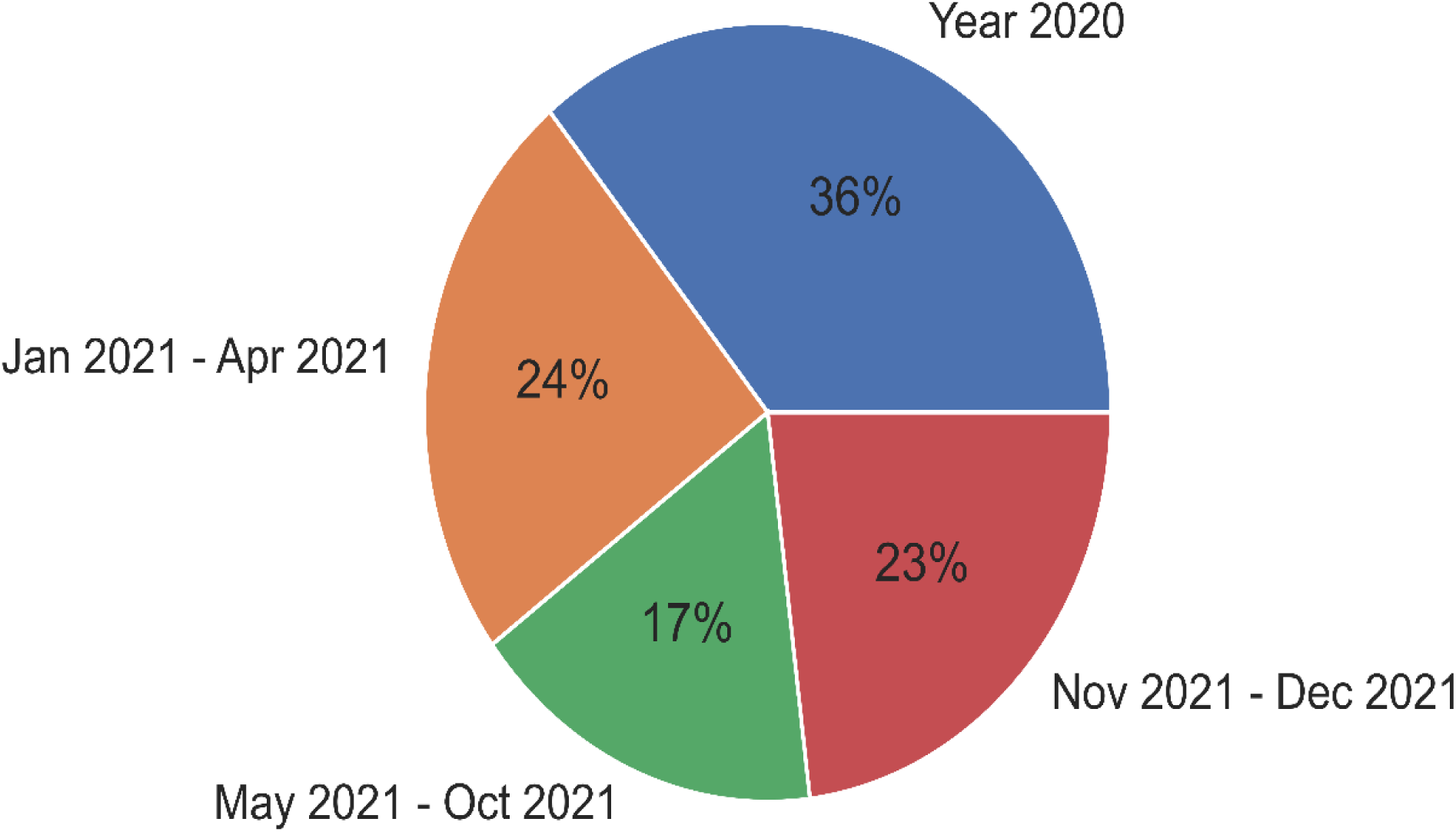
Percentages of samples collected in accordance to COVID-19 pandemic waves. (Year 2020 – mostly wild type variant, Jan 2021 to Apr 2021 – mostly beta variant, May 2021 to October 2021 – mostly delta variant and Nov 2021 to Dec 2021 – mostly omicron variant). This pie chart is necessary to relate observed trends in the SARS-CoV-2 lineages distribution where some variants might have been underestimated due to lower sample collections during their time of predominance.

### 4.2: Recombination Analyses

No evidence of recombination signals was picked by all Recombination methods in the analyses.

### 4.3: Substitution Analyses

#### 4.3.1: Phylogenetic Tree

**Figure 3:**
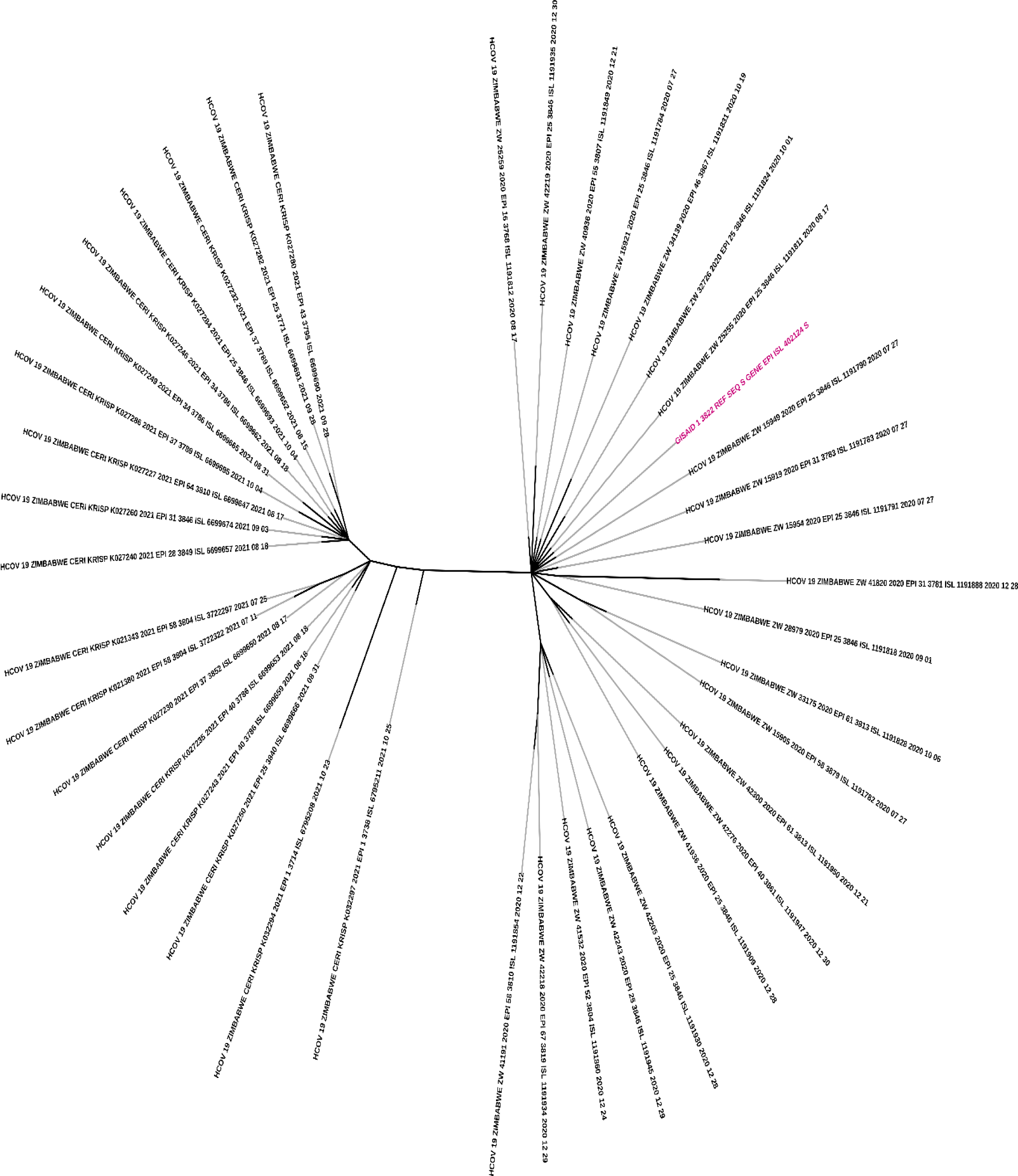
Fitted Tree based on the FUBAR Analysis results. The GISAID Reference S gene sequence (in red) was used as a central point with which other sequences diverged from. S gene sequences for samples collected in 2020 are clustering together with the GISAID Reference S gene sequence whereas samples collected in 2021 are clustering away from the GISAID Reference S gene sequence.

#### 4.3.2: Sites showing strong positive selection based on the Bayes Factor

**Figure 4:**
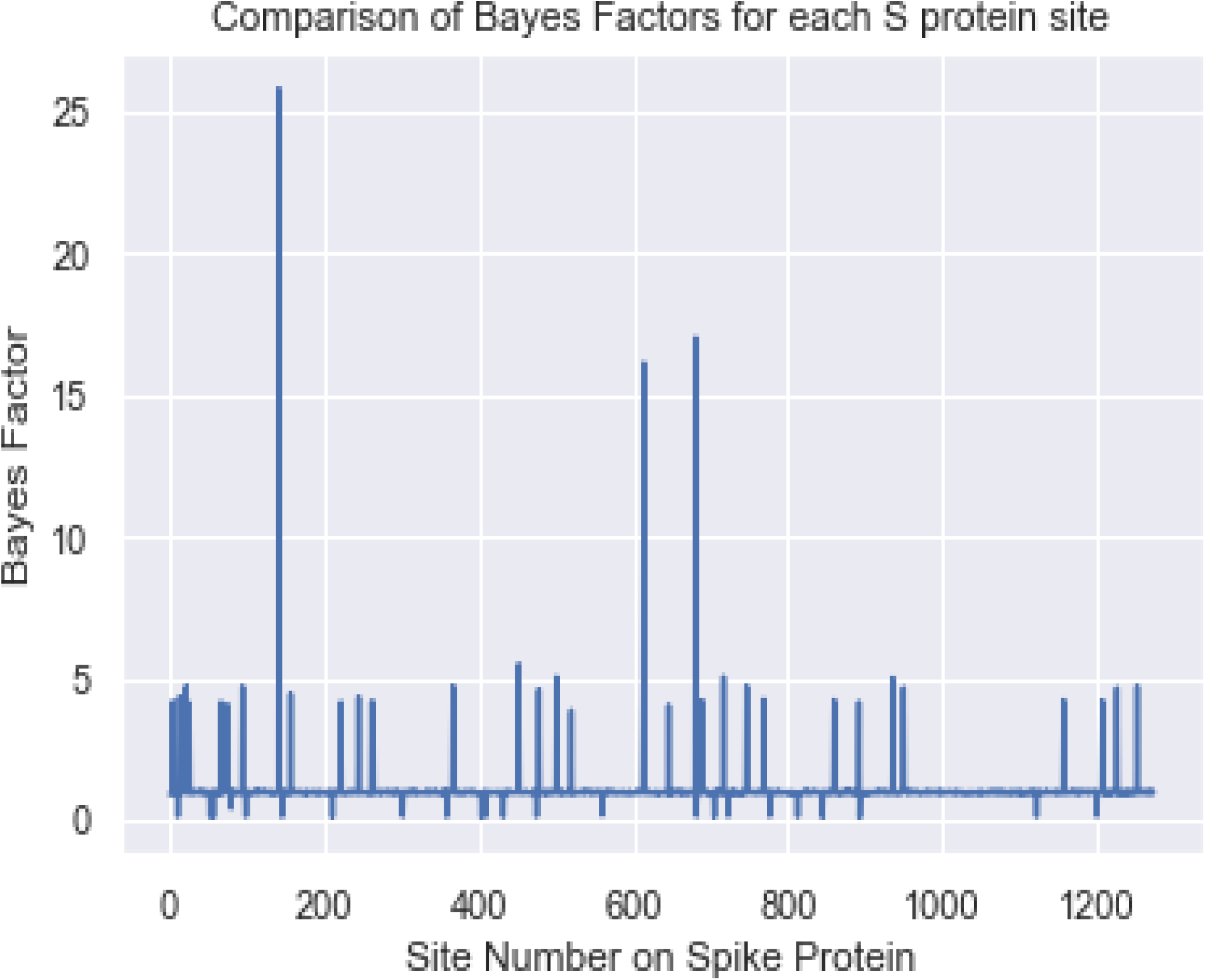
A plot investigating sites showing strong positive selection using Bayes Factor values. 3 sites in the ranges between sites 100 – 200 and sites 600 – 800 showed strong positive selection represented by the 3 outstanding peaks with Bayes Factor > 15.

#### 4.3.4: Overall summary of sites showing positive and negative selection

**Table 1:**
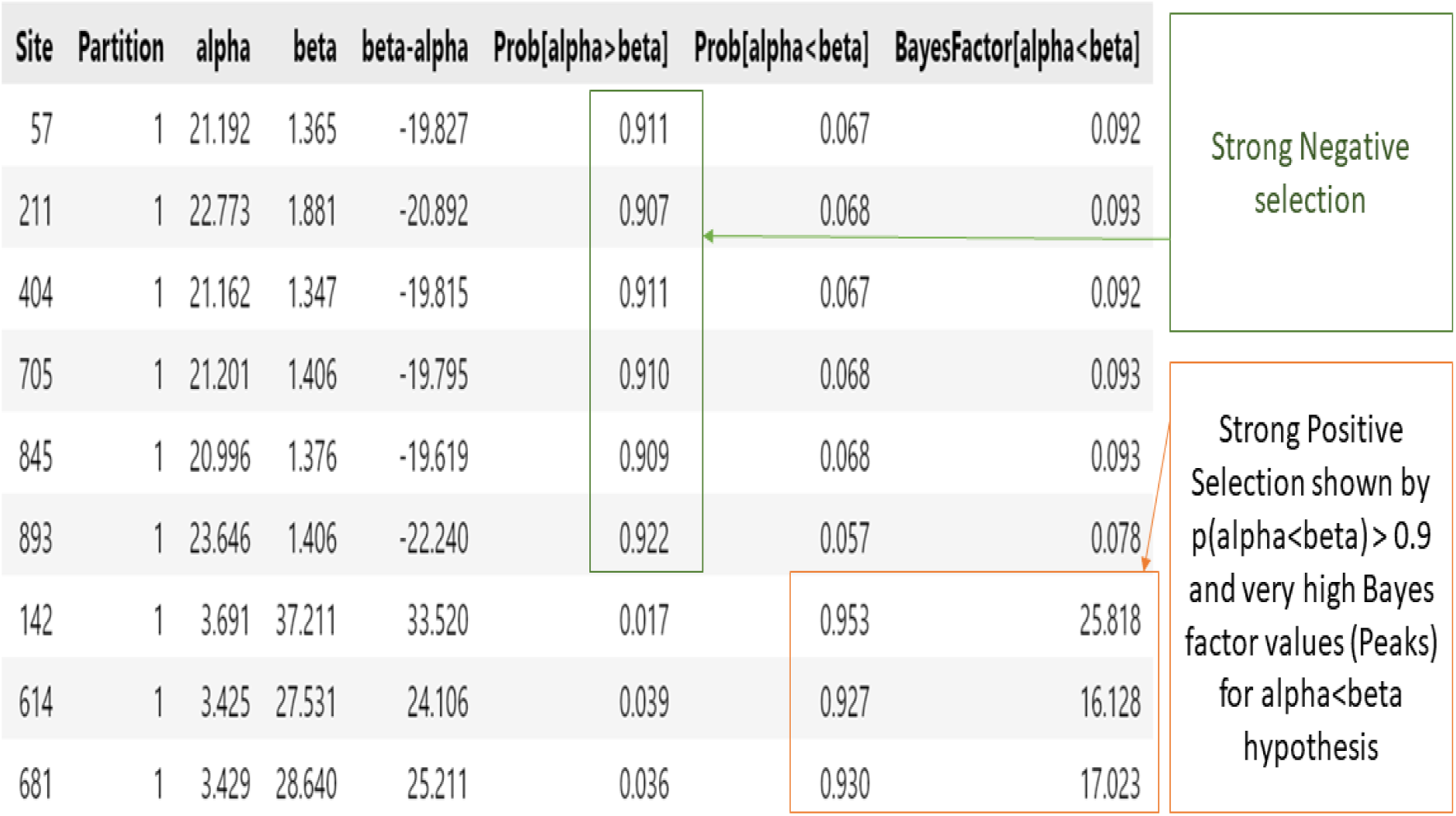
Sites that showed strong negative selection with posterior probabilities [p(α> β) > 0.9] and sites showing strong positive selection with posterior probabilities [p(β > α) > 0.9]

## 4.4: Discussion

The SARS-CoV-2 variant that was present in most of the samples collected in Zimbabwe is Beta lineage according to data with 36% coverage (Fig 1). The Beta variant caused the second COVID-19 wave in Zimbabwe however this is not to say that it was the most predominant variant thus far in Zimbabwe. The bias is addressed by Figure 2 which shows that a higher number of samples were collected during the second wave i.e. excluding 2020 samples perhaps due to availability of resources at that moment for sequencing SARS-CoV-2. The Delta variant which caused the third wave could have been underestimated because approximately 15% of SARS-CoV-2 sequences were sampled at its time of prevalence. The Omicron variant which caused the fourth wave is the most fit SARS-CoV-2 variant which contributed 23% of the COVID-19 infections in Zimbabwe outcompeting the likes of Delta variant within a space of 2 months possibly due to its multiple mutations incurred within its genome.

From the Phylogenetic tree (Figure 3), the samples collected in 2020 are clustering together with the GISAID Reference S gene sequence because at this time a few mutations had been acquired by the SARS-CoV-2. There is also a general clustering of sequences which were collected around the same time of the year on the GISAID Reference S gene sequence branch. Sequences of samples collected in 2021 diverged onto another branch away from the GISAID Ref Seq showing that at this period SARS-CoV-2 had acquired a number of mutations which resulted in this divergence. However, with 2021 samples the trend of samples collected around the same time is absent because the samples were collected at shorter intervals compared to 2020 samples resulting in overlaps of acquired mutations. The tree gives us confidence that FUBAR analysis results are relatively accurate.

The Bayes Factor from the FUBAR table of values is testing the Hypothesis for strong evidence of a site undergoing Positive selection (α> β). 3 sites showed strong evidence of positive selection represented by the outstanding peaks above 15 in the bar graph (Figure 4). To pinpoint which sites generated peaks in the above graph, the dataset was tested for sites with a threshold of Posterior probabilities > 0.9 in favour of positive selection and the mutations were identified as D142G, P681R and D614G in decreasing order of strength (Table 1 in red box). Also, sites with a Posterior Probability > 0.9 in favour of negative selection were also tested to understand the dynamics of evolutionary patterns Zimbabwean Spike gene and strong negative/purifying selection was observed at sites 893, 211, 57, 404, 705 and 845 in decreasing order of strength.

The 3 peaks generated in the Bayes Factor graph showing evidence of strong positive selection are D142G mutation in the N-terminal domain (NTD), P681R mutation in the Furin-cleavage site and D614G in the Receptor-binding domain (RBD) of the S protein. S protein site 142 contributes to the supersite epitope that binds NTD-directed neutralising antibodies. The D142G mutation shows back mutations where the mutation changes from D142 to G142 then reverts back to D142. This disturbance at the supersite epitope that binds NTD-directed neutralising antibodies reduces immune recognition leading to higher viral loads. (Shen *et al*; preprint). For antibodies targeting this site through vaccination or natural immunity, there is an increased probability of re-infection when the mutation takes place due to reduced specificity of the antibodies to the NTD. According to a preprint by Liu et al., the P681R spike mutation led to predominance of the Delta variant globally before the Omicron variant. Proline amino acid which is usually found in sites where a polypeptide chain makes sharp turns is replaced with a basic Arginine amino acid, this enhances full-cleavage of S1 and S2 proteins leading to increased infection via cell surface cell entry. The D614G has been associated with higher viral load in the upper respiratory tract but not with increased disease severity (Korber,2020). Structural analyses of the RBD revealed that the G614 form of S protein in its open conformation occupies a higher percentage of ACE2 receptor site than D614 form leading to enhanced binding to the ACE2 receptor (Yurkovitskiy, 2020). In another study the D614G exhibited efficient replication in human nasal epithelial cell lines and enhanced transmission in hACE2 transgenic mice and Syrian hamsters (Hou et al., 2020). These mutations show that the SARS-CoV-2 is a clever virus which is exploiting a number of tricks to evade and survive in the human system. By generating a consensus sequence of Zimbabwean sequences it was found that it has 99.95% identity compared to the GISAID Reference Sequence.

## 5.0: Recommendations and conclusion

SARS-CoV-2 sequences from Zimbabwean patients are showing evidence of mutations in the N-terminal domain (D142G), receptor-binding domain (D614G) and furin-cleavage site(P681R). Symptoms correlated to the acquired mutation can be used as a guide to access medicines precise for the Zimbabwean population. Use of very big datasets is recommended to reduce bias of observed mutation.

## Data Availability

All data produced in the present study are available upon reasonable request to the authors

